# PrEP-associated screening reduces *N. gonorrhoeae* transmission but increases case notifications among MSM: a modeling study

**DOI:** 10.64898/2026.06.30.26356925

**Authors:** Loïc Marrec, Sonja Lehtinen

## Abstract

The rapid scale-up of HIV pre-exposure prophylaxis (PrEP) among men who have sex with men has coincided with rising rates of bacterial sexually transmitted infections (STIs), particularly *Neisseria gonorrhoeae*. This temporal association has raised concerns that PrEP may be driving a new STI epidemic. However, the epidemiological impact of PrEP reflects a trade-off between potential behavioral risk compensation, which increases transmission risk, and intensified clinical surveillance, which shortens infection duration. Determining whether PrEP amplifies or mitigates STI transmission therefore requires understanding how these competing effects balance at the population level. To address this question, we develop a transmission model stratified by sexual activity and PrEP use, derive simple analytical conditions governing changes in prevalence, incidence, and notification rates, and evaluate these dynamics using empirically informed parameter estimates. Our analysis demonstrates that current quarterly screening guidelines are generally sufficient to reduce both the true endemic prevalence and incidence of *N. gonorrhoeae*, successfully overcompensating for plausible reductions in condom use. We also confirm and expand on previous findings that clinical notification rates may surge even as the true disease burden declines, driven by the detection of previously undiagnosed asymptomatic infections. These findings suggest a shift in focus from the potential impact of PrEP on STI transmission to the consequences of increased STI diagnoses and treatment, particularly whether greater antibiotic consumption may accelerate the spread of antimicrobial resistance in *N. gonorrhoeae*.

## Introduction

Men who have sex with men (MSM) remain disproportionately affected by HIV worldwide (UNA, 2023; European Centre for Disease Prevention and Control and World Health Organization, 2024), motivating the development and widespread adoption of pre-exposure prophylaxis (PrEP) as a prevention strategy. Following the demonstration of its efficacy in trials such as iPrEx (Grant et al., 2010), PROUD (McCormack et al., 2016), and IPERGAY (Molina et al., 2015), PrEP has been rapidly scaled up and has become a cornerstone of HIV prevention among MSM. This expansion has contributed to substantial reductions in HIV incidence, including declines of 25–40% in some high-coverage settings (Grulich et al., 2018).

However, the expanding use of PrEP has coincided with rising rates of bacterial sexually transmitted infections (STIs), particularly among MSM (Ahmed and Mkpa, 2023; Xing and Escudero, 2025). Increases in infections such as gonorrhea, caused by *Neisseria gonorrhoeae* (NG), have been documented across multiple regions, with notification rates in the EU/EEA more than doubling over the past decade (European Centre for Disease Prevention and Control, 2025). This parallel trend has sparked an ongoing clinical and epidemiological debate regarding the broader consequences of PrEP implementation (Ramchandani and Golden, 2019).

The effect of PrEP on bacterial STI transmission is not straightforward to predict. On the one hand, PrEP may lead to *risk compensation*, whereby individuals engage in more condomless sex in response to reduced perceived HIV risk (Montaño et al., 2018; Traeger et al., 2019). This may increase exposure to bacterial STIs and facilitate transmission. On the other hand, PrEP delivery is embedded within a framework of intensified clinical follow-up, including regular STI screening (Parikh et al., 2025). Earlier diagnosis and treatment shorten the duration of infectiousness and interrupt transmission chains. The net population-level impact of PrEP on bacterial STI epidemics therefore depends on the balance between these competing processes.

Understanding the impact of PrEP on STI transmission dynamic is further complicated by heterogeneity in sexual networks. STI transmission is often sustained by core groups with high rates of partner change and dense connectivity (Yorke et al., 1978). Within MSM populations, individuals with higher sexual activity may disproportionately contribute to transmission, and bisexual partnerships may create bridges to heterosexual networks. These structural features can amplify or dampen the effects of behavioral and clinical changes associated with PrEP.

The public health implications are particularly important for gonorrhea, given the rapid emergence of antimicrobial resistance in NG (World Health Organization, 2023). Rising incidence leads to increased antibiotic use, which may accelerate the selection and spread of resistant strains (Fingerhuth et al., 2016; Riou et al., 2023). Understanding how PrEP influences STI transmission dynamics is therefore critical not only for controlling infection burden but also for preserving treatment effectiveness.

Modeling studies have examined the impact of PrEP on bacterial STI dynamics. For example, using a detailed network-based model of HIV, gonorrhea, and chlamydia transmission among MSM, Jenness et al. (2017) showed that the increased screening embedded within PrEP programs could offset the effects of reduced condom use, leading to net declines in STI incidence. More recently, Müller et al. (2025) demonstrated that intensified screening may increase observed STI diagnoses even as true prevalence declines, highlighting the importance of distinguishing between notification rates and underlying transmission. Together, these studies emphasize the central role of PrEP-associated screening in shaping STI epidemiology. However, the mechanisms linking screening, transmission, prevalence, incidence, and notification rates remain difficult to disentangle in complex models, and the plausibility of specific outcomes depends on how accurately model parameters capture real-world epidemiology. This motivates the development of complementary frameworks that yield simpler epidemiological insights, are easier to parameterize, and can therefore provide an important additional perspective when informing public-health recommendations.

To complement these previous modeling efforts, we develop a mechanistic framework to evaluate how PrEP uptake shapes gonorrhea transmission among MSM. Using a deterministic compartmental model stratified by sexual activity and PrEP use, we explicitly capture the interplay between behavioral change and clinical surveillance. Beyond numerical simulations, we derive simple analytical conditions governing how PrEP-associated changes in condom use and screening influence prevalence, incidence, and notification rates. This approach provides a framework for understanding when PrEP scale-up is expected to amplify or mitigate transmission and how observed epidemiological trends relate to underlying disease dynamics.

## Model and Methods

### Host population structure

We consider a MSM population of constant size, structured into groups based on their sexual activity (Figure 1A). A fraction 1−*α* of the population belongs to a low-activity group, denoted L, while the remaining fraction *α* belongs to a high-activity group, denoted H.

**Figure 1:**
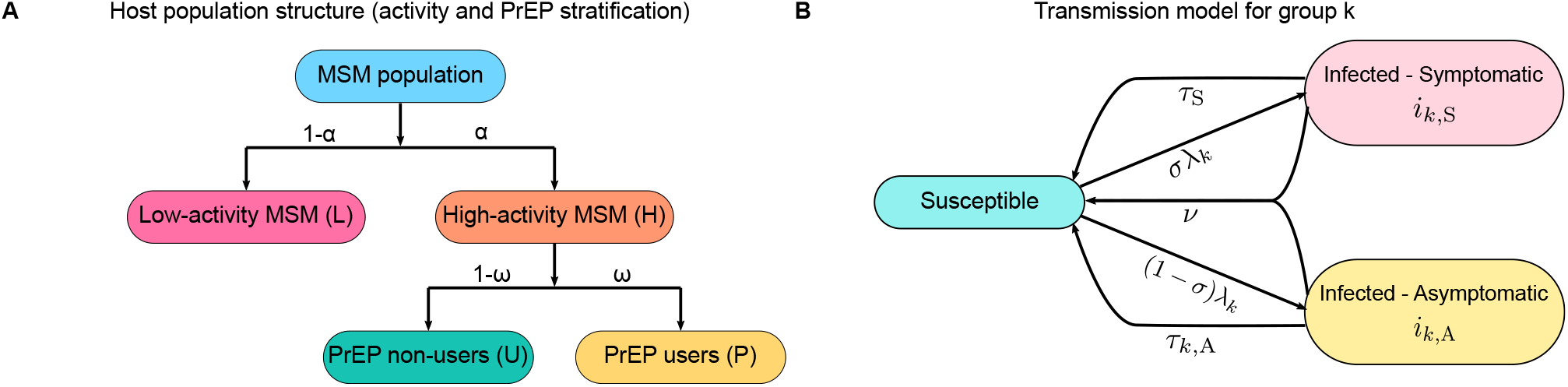
Model structure. Panel **A** shows the structure of the host population. The MSM population is divided into a low-activity group L, representing a fraction 1 − *α*, and a high-activity group H, representing a fraction *α*. The high-activity group is further subdivided into PrEP users P (fraction *ω*) and PrEP non-users U (fraction 1 − *ω*). Panel **B** illustrates the transmission dynamics. Susceptible individuals in group *k* ∈ {L, P, U} become infected at a rate determined by the force of infection, *λ*_*k*_. Upon infection, individuals develop symptoms with probability *σ* or remain asymptomatic with probability 1 − *σ*. Symptomatic individuals return to the susceptible state through treatment at rate *τ*_S_, assumed to be identical across groups, whereas asymptomatic individuals do so through testing followed by treatment at rate *τ*_*k*,A_, which may vary by group. In addition, both symptomatic and asymptomatic individuals may recover through natural (spontaneous) clearance at rate *ν*.

Within the high-activity group, individuals are further stratified according to PrEP use: a fraction *ω* uses PrEP (group P), whereas a fraction 1 − *ω* does not use PrEP (group U).

Thus, the host population is divided into three epidemiological groups: *G* = {L, P, U}. Each group *k* ∈ *G* is characterized by a partner-change rate *π*_*k*_, a transmission probability per partnership *β*_*k*_, and an asymptomatic testing rate *τ*_*k*,A_, which may differ across groups.

### Transmission model

To describe the transmission dynamics of infection in the structured MSM population, we use a deterministic approach based on a system of ordinary differential equations (ODEs). Since the host population size is assumed to be constant, we track the time evolution of the population densities of infected individuals within each host group.

For each group *k* ∈ *G*, we denote by *i*_*k*,S_(*t*) and *i*_*k*,A_(*t*) the densities of symptomatic and asymptomatic infected individuals, respectively. The density of susceptible individuals in group *k* is therefore given by

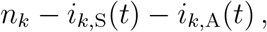

where *n*_*k*_ is the total density of the population belonging to group *k*, with the overall total population density being set to 1. In particular, we have

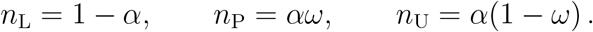

Susceptible individuals in group *k* form sexual partnerships at rate *π*_*k*_. Transmission from group *k* to *l* depends on the sexual mixing pattern between the two groups, described by the mixing matrix, whose coefficients *ρ*_*kl*_ are defined as (Supplementary Material – Section 2.1.3)

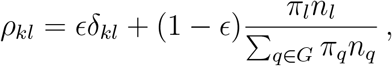

with *δ*_*kl*_ = 1 if *k* = *l* and 0 otherwise. The parameter 0 ≤ *ϵ* ≤ 1 quantifies the degree of assortative mixing, ranging from *ϵ* = 0 (proportionate mixing) to *ϵ* = 1 (fully assortative mixing, where all partnerships occur within the same group). For example, in the full proportionate mixing case, the relative transmission to each group is determined by the relative partner change rate.

Transmission also depends on the per-partnership transmission probability *β*_*kl*_. We assume that transmission probabilities between groups are symmetric and given by

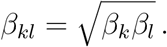

Susceptible individuals in group *k* ∈ {L, P, U} become infected at a rate determined by the force of infection, *λ*_*k*_, which satisfies (Supplementary Material − Section 2.1.2)

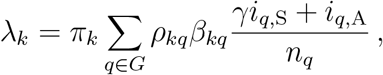

where 0 ≤ *γ* ≤ 1 represents the reduction in infectious contacts due to symptoms (*γ* = 1 no reduction; *γ* = 0 complete cessation of sexual activity).

Upon infection, individuals develop symptoms with probability *σ* or remain asymptomatic with probability 1 − *σ*. Symptomatic individuals return to the susceptible state through treatment at rate *τ*_S_, which is assumed to be identical across all groups. Asymptomatic individuals return to the susceptible state either through natural clearance at rate *ν* or through testing followed by treatment at rate *τ*_*k*,A_.

Under these assumptions, the dynamics of infection in each group *k* ∈ *G* are governed by the following system of coupled ODEs (Supplementary Material − Section 2.1.1)

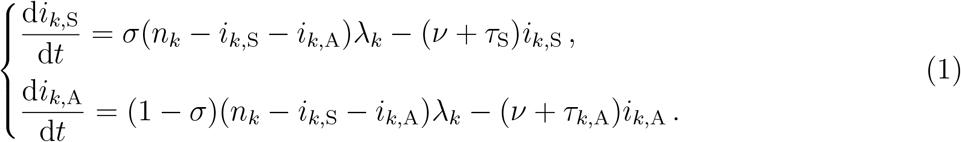

### Endemic prevalence and incidence

The two main epidemiological quantities we will consider in the following are the endemic prevalence and incidence. Prevalence refers to the total burden of infection, that is, the proportion of individuals in a population who are infected at a given point in time. By contrast, incidence measures the number of new infections that occur over a defined period of time.

For each group *k* ∈ *G*, the endemic prevalence is defined as the equilibrium values 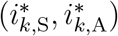 satisfying d*i*_*k*,S_*/*d*t* = 0 and d*i*_*k*,A_*/*d*t* = 0 in System 1. In general, System 1 may admit multiple equilibria, including the trivial disease-free equilibrium 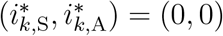. In this work, we will focus on stable equilibria, which determine the long-term epidemiological outcomes.

Incidence is defined as the number of new (or equivalently, resolved) infections per unit time at equilibrium. At endemic equilibrium, incidence equals the total outflow from the infected compartments. Hence, for each group *k*, it is obtained by multiplying the endemic prevalence by the corresponding removal rates, namely 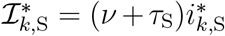 for symptomatic individuals and 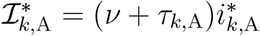 for asymptomatic individuals.

To gain further analytical insight into the model dynamics, we will derive explicit expressions for the endemic prevalence and incidence under the assumption of fully assortative mixing (*ϵ* = 1). In this limiting case, individuals form partnerships exclusively within their own group, so that each group becomes epidemiologically isolated from the others. Although the empirical mixing patterns among MSM are not perfectly assortative, our model indicates a strong tendency toward assortative partnership formation (see Model calibration), making this approximation both analytically tractable and epidemiologically informative. The resulting analytical predictions help clarify the mechanisms governing endemic behavior and provide useful benchmarks for interpreting the full model. Detailed derivations are presented in Supplementary Material – Section 3.

### Model calibration

Model parameters were informed by published demographic literature and calibrated using a rejection-based Approximate Bayesian Computation (ABC) approach. As summarized in Table 1, parameters are grouped into population-specific, group-specific, and disease-specific categories. Detailed prior distributions, calibration targets, and posterior estimates are provided in the Supplementary Material – Section 1.

**Table 1:**
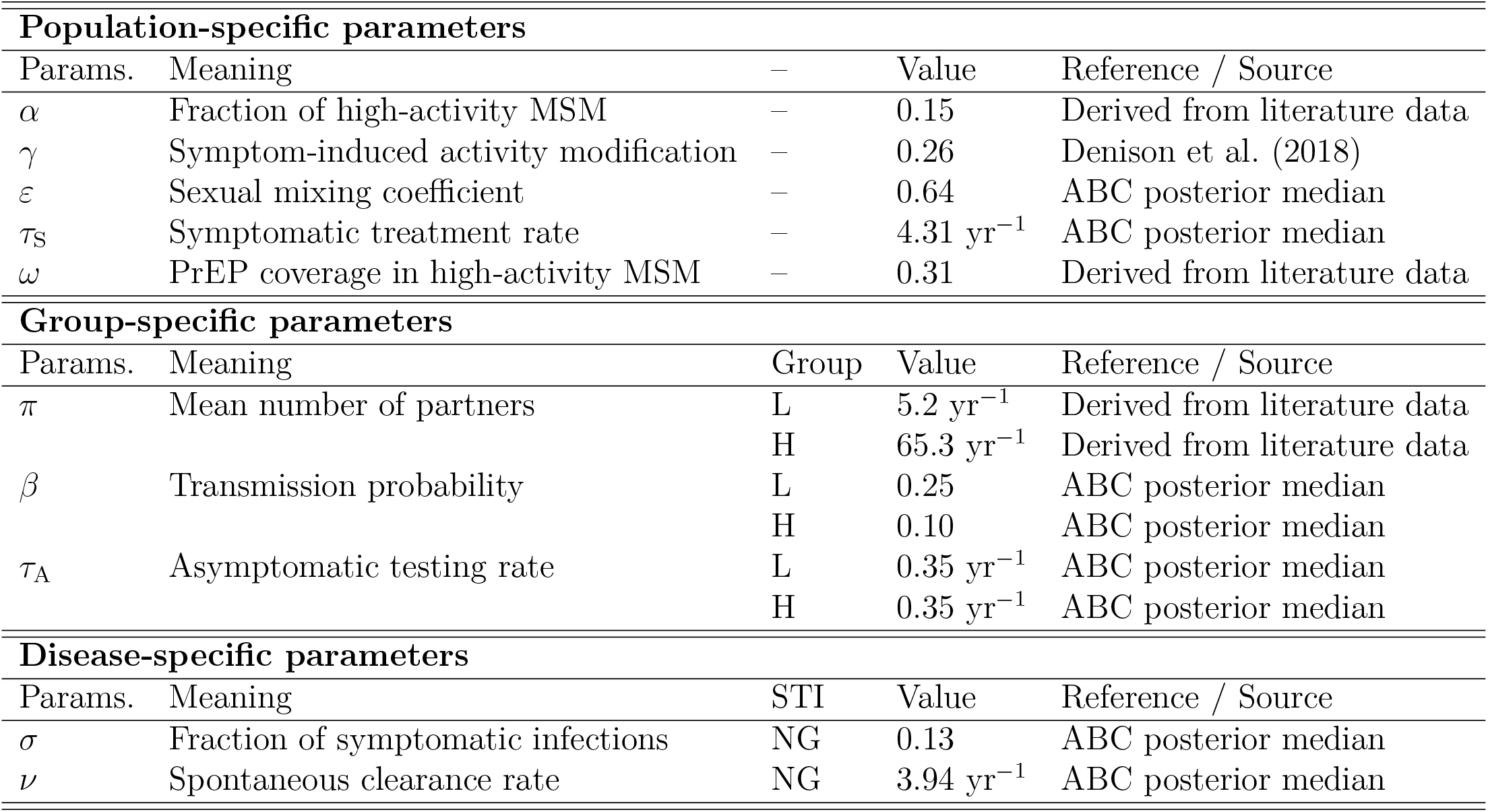
Model parameters. The table reports parameter values used in the transmission model, distinguished by population-specific, group-specific, and disease-specific characteristics. L and H stand for low- and high-activity, respectively, while NG denotes *Neisseria gonorrhoeae*. Values reported for H serve as baseline values for both high-activity PrEP non-users (U) and PrEP users (P): parameters for U are identical to H (*τ*_U,A_ = *τ*_H,A_, *β*_U_ = *β*_H_), while parameters for P are defined relative to H: *τ*_P,A_ ≥ *τ*_H,A_ and *β*_P_ ≥ *β*_H_. See Supplementary Material − Sections 1 and 2 for more detail.

| Population-specific parameters |  |  |  |  |
| --- | --- | --- | --- | --- |
| Params. | Meaning | – | Value | Reference / Source |
| $\alpha$ | Fraction of high-activity MSM | – | 0.15 | Derived from literature data |
| $\gamma$ | Symptom-induced activity modification | – | 0.26 | Denison et al. (2018) |
| $\varepsilon$ | Sexual mixing coefficient | – | 0.64 | ABC posterior median |
| $\tau_S$ | Symptomatic treatment rate | – | 4.31 yr<sup>−1</sup> | ABC posterior median |
| $\omega$ | PrEP coverage in high-activity MSM | – | 0.31 | Derived from literature data |
| Group-specific parameters |  |  |  |  |
| Params. | Meaning | Group | Value | Reference / Source |
| $\pi$ | Mean number of partners | L | 5.2 yr<sup>−1</sup> | Derived from literature data |
|  |  | H | 65.3 yr<sup>−1</sup> | Derived from literature data |
| $\beta$ | Transmission probability | L | 0.25 | ABC posterior median |
|  |  | H | 0.10 | ABC posterior median |
| $\tau_A$ | Asymptomatic testing rate | L | 0.35 yr<sup>−1</sup> | ABC posterior median |
|  |  | H | 0.35 yr<sup>−1</sup> | ABC posterior median |
| Disease-specific parameters |  |  |  |  |
| Params. | Meaning | STI | Value | Reference / Source |
| $\sigma$ | Fraction of symptomatic infections | NG | 0.13 | ABC posterior median |
| $\nu$ | Spontaneous clearance rate | NG | 3.94 yr<sup>−1</sup> | ABC posterior median |

Population-specific parameters describe the structure of the MSM population and patterns of sexual mixing. Individuals were stratified into low- and high-activity groups using a threshold of 20 sexual partners per year. This threshold was chosen to yield a realistic high-activity group size, with approximately 15% of individuals classified as high activity (*α* = 0.15), consistent with the notion of a relatively small core group contributing disproportionately to transmission (Yorke et al., 1978). To ensure our conclusions do not depend on this specific classification, we tested alternative cutoffs (*>* 10 and *>* 50 partners per year) in a sensitivity analysis, which confirmed that our qualitative results are highly robust (Supplementary Material – Section 5.1). This estimate was based on a secondary analysis of international behavioral surveys, adjusted to account for sexually inactive individuals (Mendez-Lopez et al., 2022). The degree of assortative mixing, *ϵ*, was estimated via our ABC framework, yielding a posterior median of 0.60. This indicates a relatively high degree of preferential within-group contact, consistent with previous literature (Fingerhuth et al., 2016). The proportion of PrEP users within the high-activity group, *ω*, was fixed at 0.31 to reflect plausible coverage levels (see Supplementary Material – Section 2).

Group-specific parameters govern contact rates and transmission probabilities. The mean number of partners per year, *π*, was derived directly from Mendez-Lopez et al. (2022), resulting in *π*_L_ = 5.2 year^−1^ for the low-activity group and *π*_H_ = 65.3 year^−1^ for the high-activity group. Baseline per-partnership transmission probabilities, *β*, were calibrated via ABC, yielding posterior medians of *β*_L_ = 0.25 and *β*_H_ = 0.10. Consistent with previous modeling work (Fingerhuth et al., 2016), we assumed that individuals in the high-activity group have fewer sex acts per partnership on average, resulting in a lower per-partnership transmission probability despite a higher rate of partner change. We assume that PrEP use does not affect the mean number of partners, but that it may influence transmission risk through changes in condom use. We therefore allow the transmission probability among PrEP users to differ from that of non-users, such that *β*_P_ ≥ *β*_U_. Although we explore a range of values for *β*_P_, we consider *β*_P_ = 0.15 as a reference value, corresponding to a 50% increase in transmission probability relative to *β*_U_ = 0.10 and intended to represent PrEP-associated reductions in condom use (Montaño et al., 2018).

Disease-specific parameters govern the clinical course of infection, testing, and treatment. The fraction of symptomatic infections, *σ*, and the spontaneous clearance rate, *ν*, were calibrated via ABC. A non-informative uniform prior was used for *σ*, whereas *ν* was assigned a literature-informed Gamma prior centered on 3.80 year^−1^. The resulting posterior medians were 0.13 and 2.99 year^−1^, respectively. The symptomatic treatment rate, *τ*_S_, and asymptomatic testing rate, *τ*_A_, were similarly estimated using priors informed by empirical studies (Denison et al., 2018; Jenness et al., 2017), centered on 9.33 year^−1^ and 0.88 year^−1^, respectively. The resulting posterior medians were 4.49 year^−1^ and 0.31 year^−1^. To capture intensified screening among PrEP users, we allow *τ*_P,A_ ≥ *τ*_U,A_, and explore a range of values for *τ*_P,A_. Current clinical guidelines recommend approximately four tests per year (Parikh et al., 2025), corresponding to *τ*_P,A_ = 4 year^−1^.

High-activity individuals not using PrEP (group U) share the same parameter values as the baseline high-activity group (H). PrEP users (group P) follow the same parameterization, except for potential differences in testing behavior and transmission probability.

### Data Availability

All numerical analyses were performed using *Mathematica* 13 (Wolfram Research, Inc.). Annotated code is publicly available at https://github.com/LcMrc and will be archived on Zenodo upon acceptance of the manuscript.

## Results

### Testing strategies in PrEP users compensate for behavioral changes and reduce prevalence and incidence

To assess the impact of PrEP-induced changes on prevalence and incidence, we solve System 1 and compute endemic prevalence and incidence across a range of values for *τ*_PA_ and *β*_P_, representing changes in asymptomatic testing (*τ*_PA_ ≥ *τ*_PU_) and transmission probability ( *β*_P_ ≥ *β*_U_), respectively.

Figure 2A shows the resulting prevalence, while Figure 2B shows the corresponding incidence. Both heatmaps display regions of relative increase (red) and decrease (blue), indicating that outcomes depend strongly on the combination of these two parameters. These patterns reflect a trade-off between two competing timescales: the time to new infection and the duration of infection. Increasing transmission probability shortens the time to new infection, tending to increase disease burden, whereas intensified screening shortens the duration of infection, tending to reduce it. Thus, as expected, increasing transmission probability raises both prevalence and incidence, whereas increasing the asymptomatic testing rate reduces them.

**Figure 2:**
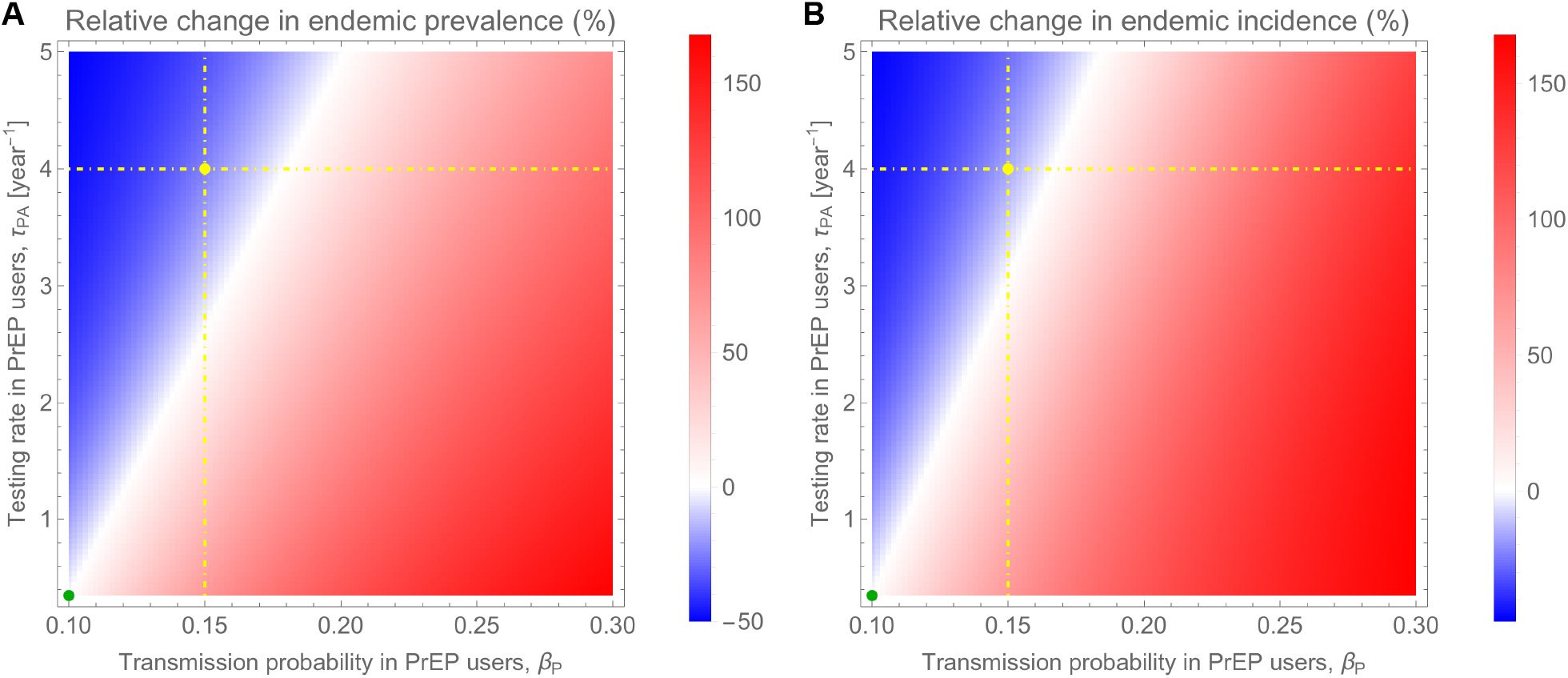
PrEP-induced changes in prevalence and incidence. Panel **A** (resp. **B**) shows the relative change in endemic prevalence (resp. incidence) of *Neisseria gonorrhoeae* as functions of *τ*_PA_ and *β*_P_. Blue (resp. red) indicates a decrease (resp. increase), and white no change. The horizontal dot-dashed line corresponds to *τ*_P,A_ = 4 year^−1^, while the vertical dot-dashed line shows *β*_P_ = 0.15. The green point represents the baseline with no behavioral change or differential testing, such that high-activity PrEP users (P) have identical parameter values to high-activity non-users (U). Other parameters are given in Table 1.

Current guidelines recommend approximately four tests per year (Parikh et al., 2025), corresponding to *τ*_P,A_ = 4 year^−1^. We further assume an increase in transmission probability to *β*_P_ = 0.15, corresponding to a 50% increase in transmission risk and intended to represent PrEP-associated reductions in condom use (Montaño et al., 2018). These values are indicated by the dot-dashed lines in Figure 2. Under these assumptions, PrEP-induced changes reduce both prevalence and incidence.

To systematically verify the robustness of this finding to parameter uncertainty, we computed the maximal percentage increase in transmission probability that could be compensated by quarterly testing (i.e. resulting in no overall increase in incidence/prevalence), across all accepted parameter sets from the ABC calibration. This sensitivity analysis confirmed that PrEP would generally require a substantial increase in transmission probability to drive an actual rise in prevalence and incidence under quarterly screening. Although a small fraction of parameter sets produced lower thresholds (5th percentile: 18.5% for prevalence and 17.5% for incidence), the median thresholds were 68.4% and 61.2%, respectively (Supplementary Material – Section 5.2).

### Incidence is less sensitive than prevalence to testing strategies in PrEP users

The parameter space in which PrEP increases disease burden is larger for incidence (Figure 2B) than for prevalence (Figure 2A). Equivalently, Figures 3 show that a higher testing frequency is required to reduce incidence than to reduce prevalence. To understand why, let us consider the system at endemic equilibrium, where the rate of new infections balances the rate at which infections are cleared.

**Figure 3:**
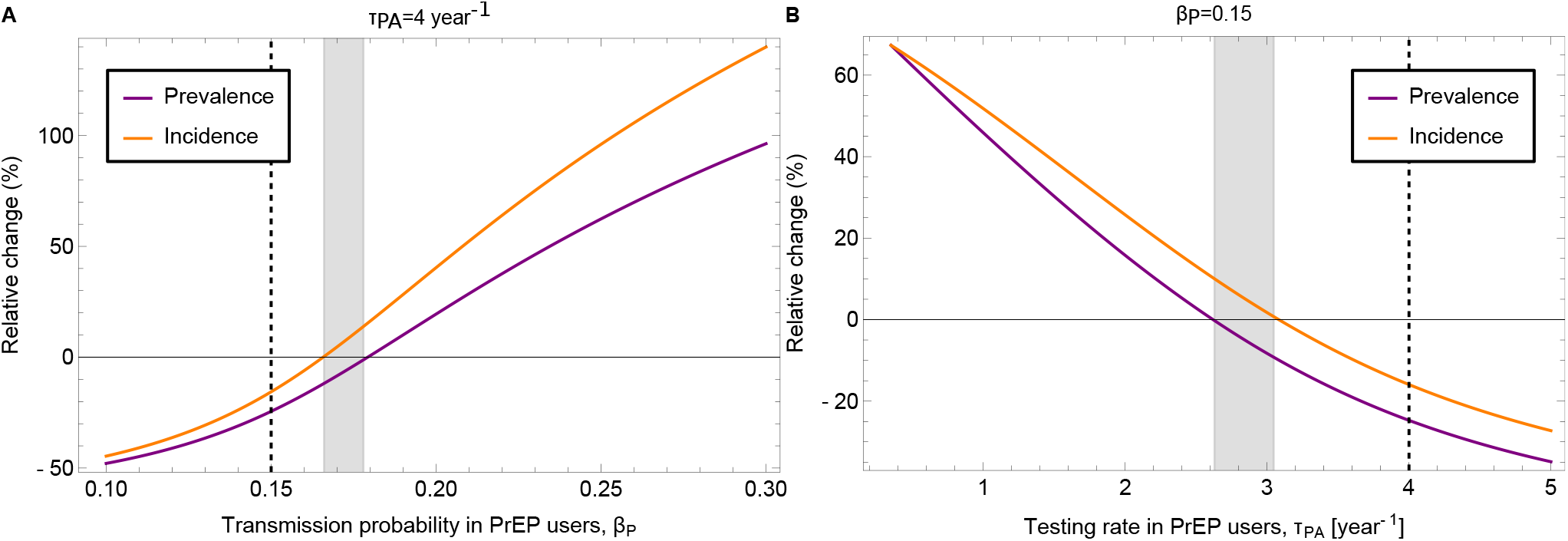
Sensitivity of prevalence and incidence to PrEP-induced changes. Panels **A** and **B** show the relative change in endemic prevalence (purple) and incidence (orange) of *Neisseria gonorrhoeae* as functions of *β*_P_ and *τ*_P,A_, respectively. In panel **A**, the testing rate is fixed at *τ*_P,A_ = 4 year^−1^, and the vertical dashed line indicates *β*_P_ = 0.15. In panel **B**, the transmission probability is fixed at *β*_P_ = 0.15, and the vertical dashed line indicates *τ*_P,A_ = 4 year^−1^. The gray shaded areas in both panels highlight the parameter space in which the relative change in prevalence is decreased while that of incidence is increased. Other parameters are given in Table 1.

Because increasing the screening frequency shortens the duration of infection, it reduces the average time individuals remain infectious and therefore lowers endemic prevalence.

Incidence behaves differently. At endemic equilibrium, it satisfies

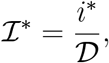

where *D* is the mean duration of infection. Increasing the screening frequency reduces prevalence and therefore tends to decrease incidence. However, it simultaneously shortens the duration of infection, which appears in the denominator of ℐ^∗^. This second effect acts in the opposite direction, partially offsetting the reduction arising from the smaller infected reservoir. Consequently, testing influences incidence through two competing pathways, whereas its effect on prevalence is purely direct. As a result, prevalence responds more strongly to increases in screening intensity than incidence, explaining why substantially higher testing frequencies are required to reduce both prevalence and incidence. In fact, incidence may even increase with more frequent testing when endemic prevalence exceeds 50%, although this regime is not relevant for gonorrhea in MSM populations (see Supplementary Material – Section 4).

This dynamic is explicitly visible in the fully assortative mixing case (*ϵ* = 1; Supplementary Material – Section 2.2), where groups are epidemiologically isolated. PrEP leads to an increase in prevalence if the basic reproduction numbers satisfy *R*_0,P_ *> R*_0,U_, or equivalently (Supplementary Material – Section 2.2.2)

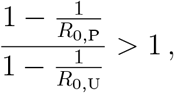

where

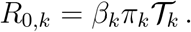

The infectious duration *T*_*k*_ is given by

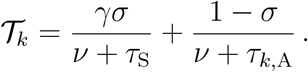

However, PrEP leads to an increase in incidence if (Supplementary Material – Section 2.2.3)

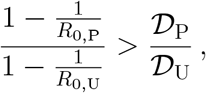

where

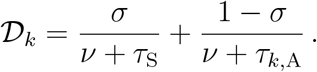

Note that the infectious duration *T*_*k*_ represents the effective time an individual can transmit the pathogen to others, whereas the infection duration *D*_*k*_ is the total time an individual remains infected. Because individuals may reduce their sexual activity upon developing symptoms (*γ* ≤ 1), the infectious duration is strictly shorter than or equal to the total infection duration (*T*_*k*_ ≤ *D*_*k*_).

Because high-frequency testing makes the ratio *D*_P_*/D*_U_ less than 1, the condition for incidence to increase is met much more easily than the condition for prevalence.

### Observed epidemiological indicators can misrepresent the true impact of PrEP on STI spread

In practice, key epidemiological quantities such as true prevalence and incidence are rarely directly observed. Prevalence is particularly difficult to measure because it requires frequent, unbiased community-wide screening to capture the large pool of asymptomatic individuals who may never seek care. Consequently, surveillance systems generally rely on the notification rate, which captures only the diagnosed fraction of true incidence, missing undiagnosed infections that clear naturally.

Figure 4 shows that resulting notification rate exhibits an increase following PrEP introduction across the vast majority of the explored parameter space. This pattern is consistent with the *testing paradox* previously described by Müller et al. (2025) for chlamydia transmission, whereby intensified screening increases case detection even as underlying transmission declines. Similarly, our results show that for NG, PrEP roll-out may lead to a rise in reported cases despite simultaneous reductions in both true prevalence and incidence (Figure 2).

**Figure 4:**
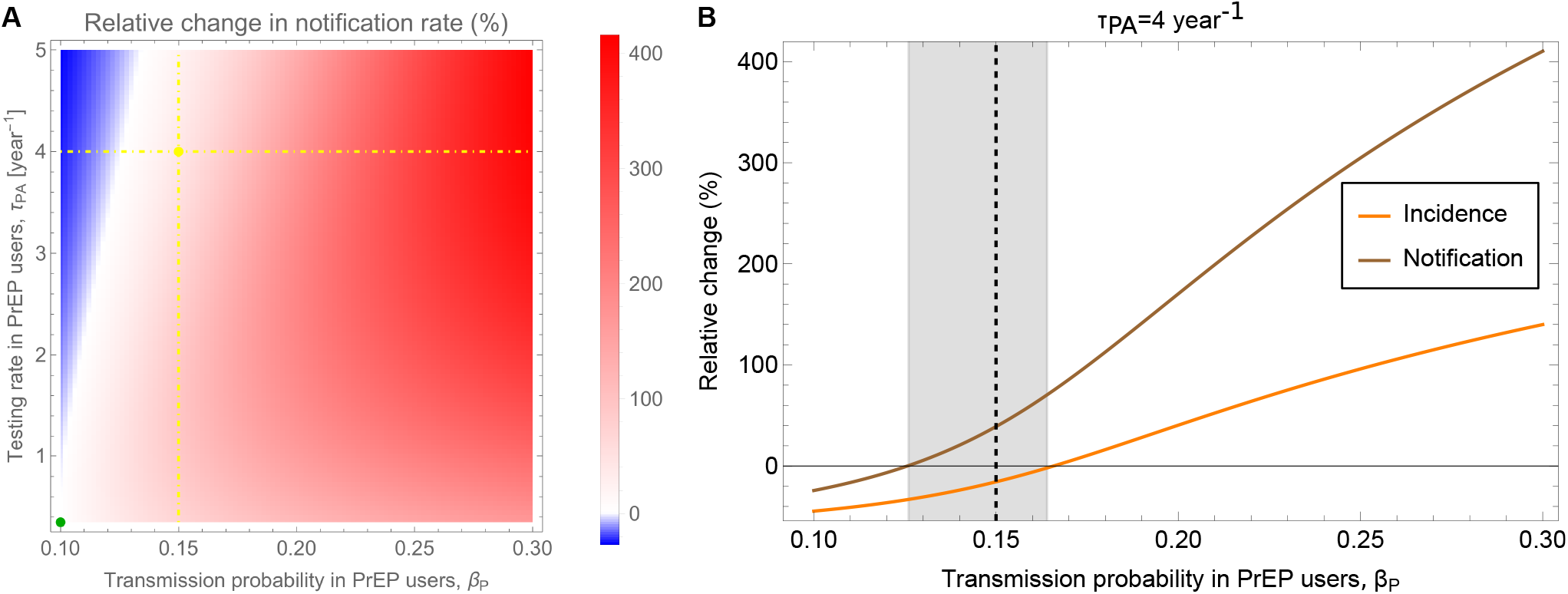
PrEP-induced changes in notification rate. Panel **A** shows the relative change in notification rate of *Neisseria gonorrhoeae* as functions of *τ*_PA_ and *β*_P_. Blue (resp. red) indicates a decrease (resp. increase), and white no change. The horizontal dot-dashed line corresponds to *τ*_P,A_ = 4 year^−1^, while the vertical dot-dashed line shows *β*_P_ = 0.15. The green point represents the baseline with no behavioral change or differential testing, such that high-activity PrEP users (P) have identical parameter values to high-activity non-users (U). Panel **B** shows the relative change in endemic incidence (orange) and notification (brown) of *Neisseria gonorrhoeae* as a function of *τ*_P,A_. The testing rate is fixed at *τ*_P,A_ = 4 year^−1^, and the vertical dashed line indicates *β*_P_ = 0.15. The gray shaded area highlights the parameter space in which the relative change in incidence is decreased while that of notification is increased. For both panels, other parameters are given in Table 1.

We can understand this by extending the logic developed for incidence. The total notification rate, *N*^∗^, is given by the product of the true incidence, ℐ^∗^, and the probability that an infection is detected before natural clearance, *ϕ*. Substituting our equilibrium relationship for incidence (ℐ^∗^ = *i*^∗^*/D*), the notification rate is rewritten as

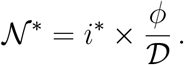

Increasing the screening frequency reduces prevalence and therefore tends to decrease notifications by shrinking the infected reservoir. As discussed above, it also shortens the duration of infection, which tends to offset the reduction arising from the smaller infected reservoir. In addition, more frequent screening raises the probability that an infection is detected before natural clearance. Consequently, testing influences the notification rate through three competing pathways: a reduction in prevalence, a shortening of infection duration, and an increase in case detection. The latter two effects act in the same direction and can outweigh the decline in prevalence, causing notification rates to rise even as the underlying burden of infection decreases.

This effect is explicitly visible in the fully assortative case (*ϵ* = 1; Supplementary Material – Section 2.2). PrEP-associated changes induce an increase in the notification rate if (Supplementary Material – Section 2.2.3)

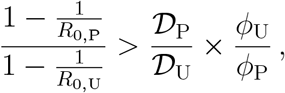

where

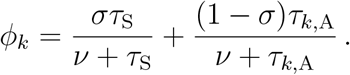

Because PrEP users are tested more frequently than non-users (*ϕ*_P_ > *ϕ*_U_), the ratio *ϕ*_U_*/ϕ*_P_ is less than 1. This multiplier further lowers the threshold required to trigger an increase in notifications compared to the threshold for incidence.

To systematically verify the robustness of this finding to parameter uncertainty, we computed the increase in transmission probability required to cause a net increase in notifications across all accepted parameter sets from the ABC calibration, for quarterly testing. A modest increase in transmission probability was sufficient for PrEP to induce an increase in case notifications. The median transmission threshold was 36.0%, and the 5th percentile was only 3.6%, indicating that increases in notifications occurred under relatively small transmission increases for a subset of parameter sets (Supplementary Material – Section 5.2).

## Discussion

The rapid scale-up of PrEP has revolutionized HIV prevention among MSM, yet its collateral impact on bacterial STI dynamics remains a subject of clinical and epidemiological debate. While concerns regarding potential *risk compensation*, specifically increased condomless sex, are prevalent (Montaño et al., 2018; Traeger et al., 2019), these behavioral shifts occur within a framework of intensified clinical surveillance (Parikh et al., 2025). Our results provide a mechanistic evaluation of this trade-off and suggest that, under current clinical recommendations, PrEP-associated screening is likely to outweigh the transmission-promoting effects of plausible reductions in condom use.

Specifically, our model predicts that quarterly screening among PrEP users is generally sufficient to offset a 50% increase in transmission probability among PrEP users and, thus, reduce both the endemic prevalence and incidence of NG (Figures 2A and B). Sensitivity analyses confirm that these conclusions are highly robust to alternative assumptions regarding the network structure, such as the threshold used to define the high-activity group, and hold true across the posterior distribution of calibrated parameters (Supplementary Material – Section 5). These findings are consistent with previous modeling work suggesting that frequent STI screening can compensate for behavioral changes associated with PrEP use (Jenness et al., 2017). Taken together, these results suggest that the observed rise in NG notifications following PrEP roll-out should not be interpreted as evidence that PrEP is driving a new STI epidemic. Rather, under realistic assumptions, PrEP programs are expected to reduce transmission while simultaneously increasing case detection.

Our analysis nevertheless highlights an important distinction between prevalence and incidence. Increasing testing directly reduces prevalence by shortening the duration of infection and thereby lowering the effective reproduction number. Incidence is also reduced, but less strongly, because it depends not only on the number of infected individuals but also on the rate at which infections are cleared. Consequently, prevalence responds more readily to increases in screening intensity than incidence, and higher testing frequencies are required to achieve comparable reductions in the number of new infections. This differential sensitivity provides a simple mechanistic explanation for why prevalence may decline substantially even when incidence changes only modestly.

Our results also connect naturally to the *testing paradox* described by Müller et al. (2025). Using a model of chlamydia transmission, these authors showed that intensified screening can increase observed case counts even while reducing the true burden of infection. We find a similar phenomenon for NG, but further show that testing affects prevalence, incidence, and notifications differently. Whereas prevalence is most sensitive to increased screening, incidence responds more weakly, and notification rates may increase even when both prevalence and incidence decline. This hierarchy of sensitivities helps reconcile apparently contradictory epidemiological observations and highlights the importance of distinguishing between observed and underlying measures of disease burden when evaluating PrEP programs.

This finding has important implications for antimicrobial resistance. Even in scenarios where PrEP-associated screening successfully reduces prevalence and incidence, notification rates often increase because a larger fraction of infections are detected and treated. For NG, this implies greater antibiotic consumption despite improvements in transmission control. Given the growing threat of antimicrobial resistance in gonorrhea (Wi et al., 2017; World Health Organization, 2023; Maatouk et al., 2025), the central public-health question may therefore be shifting from whether PrEP promotes STI transmission to whether intensified screening and treatment programs increase antibiotic exposure sufficiently to influence resistance evolution (Tsoumanis et al., 2023). Addressing this question will require integrating transmission dynamics with models of antimicrobial resistance emergence and spread (Fingerhuth et al., 2016; Riou et al., 2023).

In conclusion, our study suggests that PrEP-associated screening is likely to reduce NG transmission despite plausible reductions in condom use. At the same time, increased testing fundamentally alters the relationship between prevalence, incidence, and observed case counts, making surveillance data difficult to interpret in isolation. While rising notification rates may not signal a worsening epidemic, they do imply increased treatment activity. Understanding the long-term consequences of this increased antibiotic use may therefore be one of the most important challenges for STI control in the PrEP era.

## Author Contributions

- **Conceptualization:** Loïc Marrec
- **Formal Analysis:** Loïc Marrec
- **Funding Acquisition:** Sonja Lehtinen
- **Investigation:** Loïc Marrec
- **Methodology:** Loïc Marrec, Sonja Lehtinen
- **Software:** Loïc Marrec
- **Supervision:** Sonja Lehtinen
- **Visualization:** Loïc Marrec
- **Writing – Original Draft Preparation:** Loïc Marrec
- **Writing – Review & Editing:** Loïc Marrec, Sonja Lehtinen

## Conflict of interest declaration

The authors declare no conflicts of interest.

## Funding

The study was funded by a Swiss National Science Foundation (SNSF) grant to SL. The grant number is PR00P3_201618.

## Acknowledgments

The authors thank the EE Group at Unil for insightful discussions.

## Notes

### Competing Interest Statement

The authors have declared no competing interest.

