## Supplementary Material for "PrEP-associated screening reduces *N. gonorrhoeae* transmission but increases case notifications among MSM: a modeling study"

### 1 Model calibration

#### 1.1 Approximate Bayesian computation

We calibrated the model using a rejection-based *Approximate Bayesian Computation* (ABC) approach. A total of $10^7$ parameter sets were sampled independently from the prior distributions described below. For each sampled parameter set, the deterministic transmission model was solved numerically until equilibrium.

For each successful simulation, endemic prevalence and notification rate were computed from the model outputs and compared with predefined epidemiological calibration targets. A parameter set was accepted if all simulated quantities simultaneously fell within their corresponding admissible ranges. This procedure therefore implemented a hard-threshold ABC rejection algorithm.

The collection of accepted parameter sets was interpreted as an approximate posterior distribution consistent with the calibration targets. Out of the $10^7$ sampled parameter sets, 19,009 satisfied all calibration criteria and were retained for subsequent analyses.

Posterior distributions were compared with the corresponding prior distributions and posterior summaries were computed from the retained parameter sets.

#### 1.2 Prevalence and notification rate

The epidemiological calibration targets were selected from published prevalence and surveillance estimates for MSM populations in Europe.

For the low-activity MSM group, the endemic prevalence target was based on the estimate reported by Fingerhuth et al. (2016), corresponding to a prevalence range of 0–2.79%. Although the same study also reported prevalence estimates for the high-activity MSM group, the associated interval (1.19–100%) was considered unrealistically broad for calibration purposes. We therefore used estimates from a systematic review and meta-analysis conducted by the European Centre for Disease Prevention and Control (ECDC), which reported prevalence values ranging from 7.60% to 20.98% in higher-risk MSM populations (European Centre for Disease Prevention and Control, 2024).

The prevalence range for the overall MSM population was derived as a weighted average of the prevalence in the low- and high-activity groups

$$\text{Prev} = (1 - \alpha)\text{Prev}_\text{L} + \alpha\text{Prev}_\text{H},$$

where $\alpha$ denotes the proportion of high-activity MSM in the population. Assuming $\alpha = 0.15$, the resulting admissible prevalence interval for the overall MSM population was 1.14–5.52%.

For notification rate, we used data from the ECDC Annual Epidemiological Report for 2023 on gonorrhea (European Centre for Disease Prevention and Control, 2025a). Specifically, we extracted the annual number of reported gonorrhea cases among MSM from Figure 5 for the following countries: Czechia, Denmark, Finland, Greece, the Netherlands, Norway, Portugal, Romania, Slovakia, Slovenia and Sweden. These notification counts were combined with Eurostat demographic data for males aged 15–64 years in the corresponding countries. Assuming that MSM represent 3% of the male population, a standard assumption commonly used in epidemiological studies (Marcus et al., 2013), we derived relative incidence estimates for the MSM population. Taking the minimum and maximum values across the observation period yielded an admissible notification rate interval of 0.004–0.019%. This incidence target corresponds to the overall MSM population and does not distinguish between low- and high-activity groups.

Table S1: Epidemiological calibration targets used in the ABC procedure.

| Quantity | Population | Calibration range |
|---|---|---|
| Prevalence | Low-activity MSM | 0–2.79% |
| Prevalence | High-activity MSM | 7.60–20.98% |
| Prevalence | Overall MSM | 1.14–5.52% |
| Notification rate | Overall MSM | 0.005–0.019% |

#### 1.3 Number of partners and high-activity fraction

To parameterize the sexual contact network, we stratified the modeled MSM population into two groups based on sexual activity: a low-activity group, defined as individuals having 0 to 20 partners, and a high-activity group, defined as individuals having more than 20 partners.

Data on the number of sexual partners were derived from a secondary analysis of 13 behavioral surveys across 17 countries (Mendez-Lopez et al., 2022). Because this study explicitly excluded respondents who reported zero partners from their sample statistics ($N = 55,180$ active respondents, overall mean 15.8 partners), we adjusted the reported distributions to account for sexually inactive individuals. We assumed that 10% of the overall MSM population had zero partners (for Disease Prevention et al., 2019). This yields an implied total population of $55,180/0.90 \approx 61,311$ individuals.

For the high-activity group ($> 20$ partners), the survey data reported $9,191$ individuals with a mean of 65.3 partners. Adjusting for the total implied population, the fraction of high-activity MSM in our model, denoted by $\alpha$, is calculated as

$$\alpha = \frac{9,191}{61,311} \approx 0.15\,.$$

The mean number of partners for this group remains unaffected by the inclusion of zero-partner individuals in the broader population. Thus, we set the mean partner number for the high-activity group to $\pi_{\mathrm{H}} = 65.3$ year$^{-1}$.

For the low-activity group (0–20 partners), the population fraction is the complement, $1 - \alpha = 0.85$. To determine the mean number of partners for this group, $\pi_{\mathrm{L}}$, we deduced the total number of partnerships from the active sample. The overall active sample reported approximately $55,180 \times 15.8 \approx 871,844$ total partners, of which $9,191 \times 65.3 \approx 600,172$ partners were attributable to the high-activity group. The remaining $271,672$ partners belong to the individuals with 1 to 20 partners. Dividing these remaining partners by the total number of individuals in the low-activity group (which includes both the 1–20 subgroup and the assumed 10% with zero partners, totaling $52,120$ individuals) yields

$$\pi_{\mathrm{L}} = \frac{271,672}{52,120} \approx 5.2 \text{ year}^{-1}.$$

These derived parameters, summarized in Table S2, were used to structure the contact matrix and define the sexual behavior parameters of the two activity groups.

Table S2: Population fractions and mean number of partners by sexual activity group.

| **Group** | **Partner range** | **Population fraction ($\alpha$)** | **Mean number of partners ($\pi$)** |
|---|---|---|---|
| Low-activity | 0–20 | 0.85 | 5.2 year$^{-1}$ |
| High-activity | $> 20$ | 0.15 | 65.3 year$^{-1}$ |

#### 1.4 Sexual mixing coefficient

The sexual mixing coefficient, denoted by $\epsilon$, quantifies the degree of assortative mixing. Its value ranges from 0 (proportionate mixing) to 1 (fully assortative mixing, in which all partnerships occur within the same group).

Given this bounded range, we assign $\epsilon$ a uniform prior distribution $\mathcal{U}(0, 1)$. Figure S1 shows the corresponding posterior distribution, whose median is 0.64, indicating a relatively high degree of assortative sexual mixing.

The shape of the posterior distribution of the sexual mixing coefficient, which indicates a tendency toward assortative mixing, as well as its median, are very similar to those inferred by Fingerhuth et al. (2016), who reported a median value of 0.57.

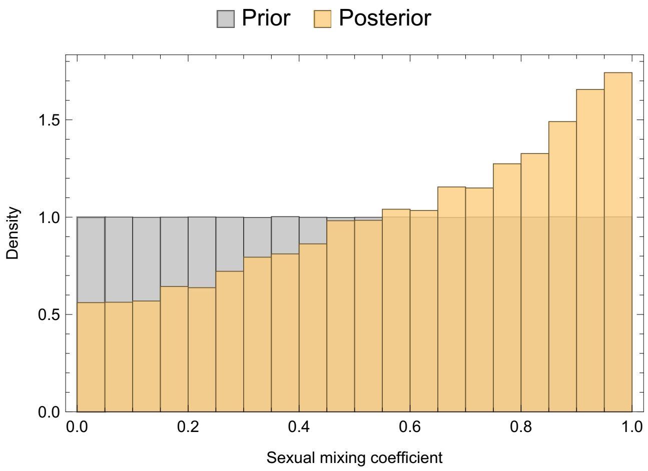

Figure S1: **Sexual mixing coefficient.** The figure shows the prior distribution of the sexual mixing coefficient, assumed to be uniform on the interval $[0, 1]$, together with its posterior distribution. The posterior median is equal to 0.64, indicating a relatively high degree of assortative sexual mixing.

#### 1.5 Transmission probability – Low-activity group

The transmission probability among low-activity MSM, denoted by $\beta_L$, quantifies the probability of transmission per partnership. Its value ranges from 0 (no transmission) to 1 (certain transmission).

Given this bounded range, we assign $\beta_L$ a uniform prior distribution $\mathcal{U}(0, 1)$. Figure S2 shows the corresponding posterior distribution, whose median is 0.25.

The shape of the posterior distribution of the transmission probability in the low-activity group is very similar to that reported by Fingerhuth et al. (2016), although they obtained a higher median value of 0.59.

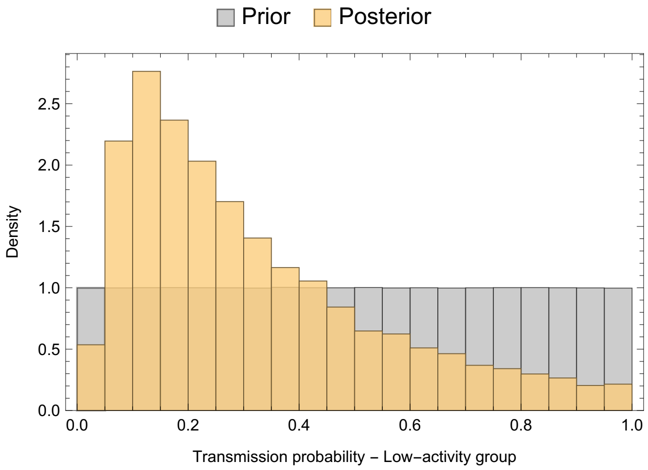

Figure S2: **Transmission probability – Low-activity group.** The figure shows the prior distribution of the transmission probability in low-activity MSM, assumed to be uniform on the interval $[0, 1]$, together with its posterior distribution. The posterior median is equal to 0.25.

#### 1.6 Transmission probability – High-activity group

The transmission probability among high-activity MSM, denoted by $\beta_\mathrm{H}$, represents the probability of transmission per partnership. Its value ranges from 0 to $\beta_\mathrm{L}$, reflecting the assumption that the effective transmission probability per partnership may be lower among high-activity MSM than among low-activity MSM.

Given this bounded range, we assign $\beta_\mathrm{H}$ a uniform prior distribution $\mathcal{U}(0, \beta_\mathrm{L})$. Figure S3 shows the corresponding posterior distribution, whose median is 0.10.

The shape of the posterior distribution of the transmission probability in the high-activity group is very similar to that reported by Fingerhuth et al. (2016), although they obtained a higher median value of 0.30.

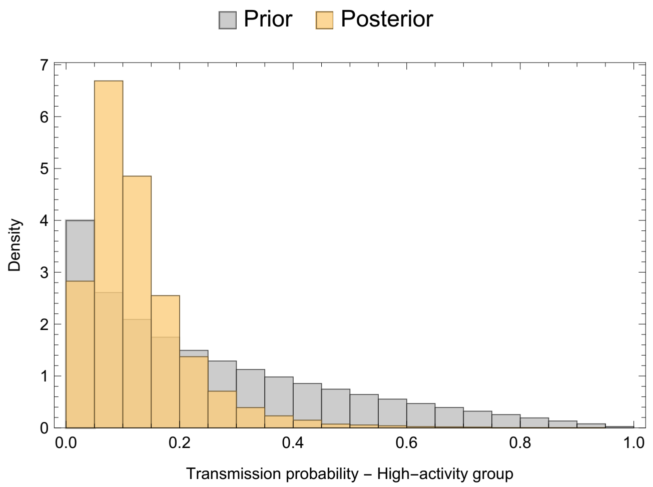

Figure S3: **Transmission probability – High-activity group.** The figure shows the prior distribution of the transmission probability in high-activity MSM, assumed to be uniform on the interval $[0, \beta_\mathrm{L}]$, together with its posterior distribution. The posterior median is equal to 0.10.

#### 1.7 Note on the transmission probability

Direct comparisons between the transmission probabilities estimated in our study and those reported in the literature should be interpreted with caution because transmission parameters are highly dependent on model structure and parameterization choices. In particular, our model does not explicitly distinguish anatomical infection sites, sexual acts, or transmission directionality. Instead, the transmission probabilities $\beta_{\mathrm{L}}$ and $\beta_{\mathrm{H}}$ represent aggregate per-partnership transmission probabilities that implicitly capture multiple behavioral and biological mechanisms.

Previous modeling studies have reported a wide range of transmission probability estimates for gonorrhea among MSM. As summarized by Jenness et al. (2017) in their supplementary material, per-sex-act transmission probabilities in male–male transmission models generally ranged from 0.20 to 0.60, with some studies reporting values as low as 0.02 and as high as 0.80. Per-partnership transmission probabilities were also highly variable, ranging from 0.10 to 0.80, depending on modeling assumptions and the anatomical sites considered.

To account for anatomical-site heterogeneity, Jenness et al. (2017) specified prior distributions of 0.30–0.60 for rectal gonorrhea transmission probability and 0.20–0.50 for urethral transmission probability. Bayesian calibration yielded posterior per-sex-act transmission probabilities of 0.36 for rectal transmission and 0.25 for urethral transmission.

Although direct comparisons are difficult because our model does not explicitly distinguish anatomical infection sites or transmission by sex act, the posterior transmission probabilities obtained here remain broadly consistent with estimates reported in previous modeling studies. Despite differences in parameter interpretation, the posterior medians obtained in our framework are of the same order of magnitude as those reported by Jenness et al. (2017).

#### 1.8 Fraction of symptomatic infections

A proportion of *Neisseria gonorrhoeae* infections are symptomatic, whereas others remain asymptomatic. We therefore define the fraction of symptomatic infections, denoted by $\sigma$, which ranges from 0 to 1.

Given this bounded range, we assign $\sigma$ a uniform prior distribution, $\mathcal{U}(0, 1)$. Figure S4 shows the corresponding posterior distribution, whose median is equal to 0.13.

Direct comparison of the estimated fraction of symptomatic infections with values reported in previous studies should be interpreted with caution due to differences in model structure and parameter aggregation. In particular, our model considers a single aggregate parameter $\sigma$ representing the overall fraction of symptomatic *Neisseria gonorrhoeae* infections in MSM, without distinguishing infection sites.

In contrast, Jenness et al. (2017) distinguish between anatomical sites and report substantially different probabilities of symptomatic infection depending on infection location. Their estimates range from approximately 0.16 for rectal infections to 0.90 for urethral infections, reflecting strong site-specific heterogeneity. When aggregated across sites and populations, their review of modeling studies reports a wide range of symptomatic proportions in men, with a central range between approximately 0.35 and 0.88, and lower estimates in some studies between 0.11 and 0.25.

Although our posterior median lies below most central estimates reported in the literature, it is broadly consistent with the lower end of reported ranges for symptomatic proportions. This difference may reflect structural aggregation in our model, which averages over anatomical sites and infection pathways, as well as differences in calibration targets and study populations.

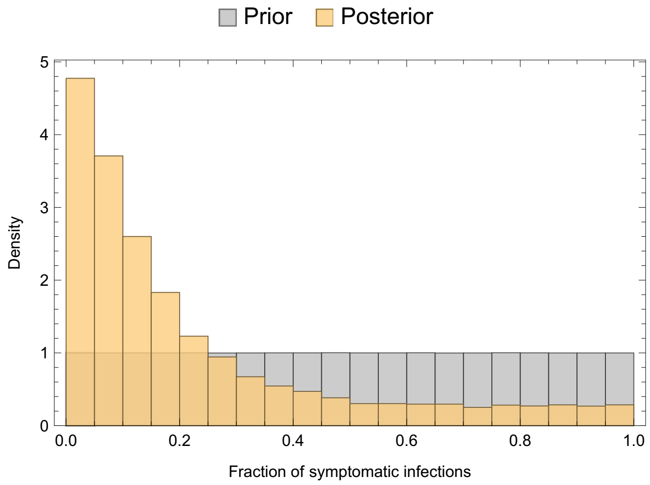

Figure S4: **Fraction of symptomatic infections.** The figure shows the prior distribution of the fraction of symptomatic infections in *Neisseria gonorrhoeae*, assumed to be uniform on the interval $[0, 1]$, together with its posterior distribution. The posterior median is equal to 0.13.

#### 1.9 Symptomatic treatment rate

Table S3: Estimation of symptomatic treatment rate from grouped duration data (Denison et al., 2018).

| Duration range | Count | Assumed value (weeks) | Weighted contribution |
|---|---|---|---|
| $\leq$ 1 week | 49 | 0.5 | 24.5 |
| 1–4 weeks | 16 | 2.5 | 40 |
| 4 weeks–6 months | 14 | 15 | 210 |
| $>$ 6 months | 5 | 39 | 195 |
| **Total** | 84 | | 469.5 |
| **Mean duration (weeks)** | | | 5.59 |
| **Mean duration (years)** | | | 0.108 |
| **Symptomatic treatment rate (year$^{-1}$)** | | | 9.33 |

Following symptom onset, individuals adopt different care-seeking behaviors: some seek treatment rapidly, while others delay considerably. We therefore estimate the symptomatic treatment rate using data from Denison et al. (2018), who analyzed healthcare-seeking behavior among individuals presenting with sexually transmitted infection symptoms at a sexual health clinic in New Zealand. Although their study did not specifically target MSM, we assume comparable behavior.

Table S3 reports the estimated mean duration of infection prior to treatment (5.59 weeks), corresponding to a treatment rate of 9.33 year$^{-1}$. This value is used as the mean of a Gamma prior distribution, $\Gamma(2, 9.33/2)$. Figure S5 shows the corresponding posterior distribution, whose median is 4.31 year$^{-1}$.

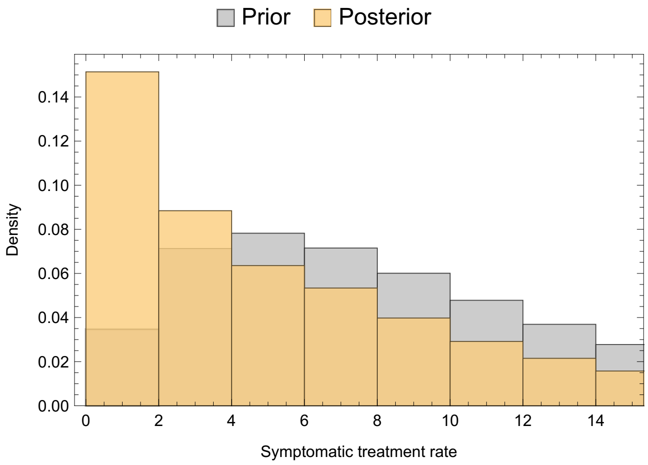

Figure S5: **Symptomatic treatment rate.** The figure shows the prior and posterior distributions of the symptomatic treatment rate, assumed to be identical across activity groups. The prior distribution is specified as a Gamma distribution, $\Gamma(2, 9.33/2)$, with mean calibrated from data reported by Denison et al. (2018). The posterior median is 4.31 year$^{-1}$.

#### 1.10 Asymptomatic testing rate

The asymptomatic testing rate, denoted by $\tau_A$, quantifies the frequency at which MSM undergo testing or screening in the absence of symptoms. Based on estimates reported by Jenness et al. (2017), who investigated STI screening practices among MSM in the United States, we assumed a mean asymptomatic testing rate of 0.88 year$^{-1}$. Because the study did not distinguish testing behavior according to sexual activity level, the same testing rate was assumed for both low- and high-activity MSM groups.

This estimate was used to parameterize the prior distribution of $\tau_A$, which was assumed to follow a Gamma distribution, $\Gamma(2, 0.88/2)$. The corresponding posterior distribution obtained after calibration is shown in Figure S6, whose median is 0.35 year$^{-1}$.

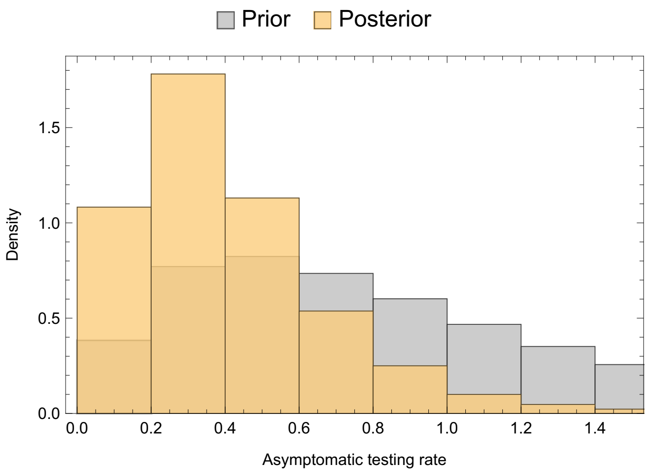

Figure S6: **Asymptomatic testing rate.** Prior and posterior distributions of the asymptomatic testing rate, assumed to be identical across activity groups. The prior distribution was specified as a Gamma distribution, $\Gamma(2, 0.88/2)$, parameterized using estimates reported by Jenness et al. (2017). The posterior median was 0.35 year$^{-1}$.

#### 1.11 Spontaneous clearance rate

Table S4: Estimation of spontaneous clearance rate for *Neisseria gonorrhoeae* infections in MSM, conditioning on asymptomatic infections.

| Site | Fraction (%) | Duration | Rate (year$^{-1}$) |
|---|---|---|---|
| Pharyngeal | 44.2 [1] | 16 weeks [2] | 3.26 |
| Rectal | 32.8 [1] | 9 weeks [3] | 5.79 |
| Urethral | 23.0 [1] | *6 months** | 2.00 |
| **Total clearance rate (year$^{-1}$)** | | | **3.80** |

Sources include: [1] Kent et al. (2005) (see Figure 2 and Table 2), [2] Barbee et al. (2021b), [3] Barbee et al. (2021a).
* Natural history studies of asymptomatic male gonorrhoea demonstrate that infections may persist for up to several months in the absence of treatment; however, to our knowledge, precise mean duration is not well characterized (Lovett and Duncan, 2019).

The spontaneous clearance rate, denoted by $\nu$, represents the rate at which untreated *Neisseria gonorrhoeae* infections resolve naturally in the absence of treatment.

To parameterize this quantity, we first constructed estimates of the mean duration of asymptomatic infection across anatomical sites. Site-specific durations for pharyngeal, rectal, and urethral infections were combined using the relative distribution of infections across sites to obtain an overall mean duration of infection. The spontaneous clearance rate was then defined as the inverse of this mean duration.

This estimate was used as the mean of the prior distribution for $\nu$, which was specified as a Gamma distribution, $\Gamma(2, 3.80/2)$. The corresponding posterior distribution obtained after calibration is shown in Figure S7, with a median value of 3.94 year$^{-1}$.

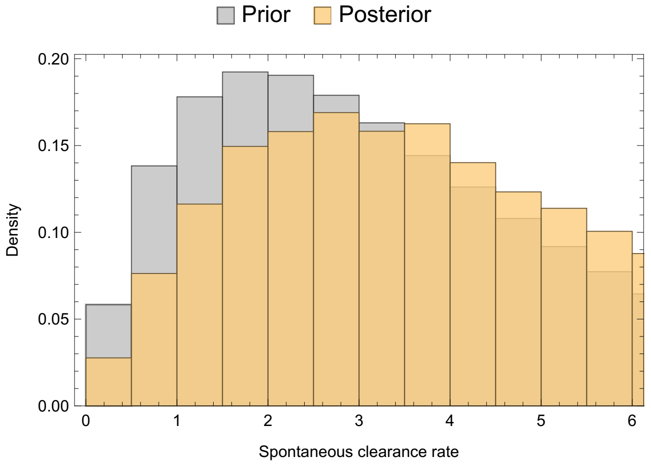

Figure S7: **Spontaneous clearance rate.** Prior and posterior distributions of the spontaneous clearance rate. The prior distribution was specified as a Gamma distribution, $\Gamma(2, 3.80/2)$. The posterior median was 3.94 year$^{-1}$.

Estimates of the duration of untreated gonorrhea infection reported in the literature exhibit substantial heterogeneity. As summarized by Jenness et al. (2017), model-based estimates of asymptomatic infection duration are often bimodal, with clusters around approximately 105–135 days and 180–185 days, while other studies report substantially shorter or longer durations ranging from a few weeks to nearly one year depending on modeling assumptions and treatment-seeking behavior.

In comparison, our calibrated posterior implies a mean infectious duration on the order of approximately 100 days, which lies within the lower range of published estimates for asymptomatic infection duration. This is broadly consistent with the lower cluster of estimates reported in the literature, although direct comparison is complicated by differences in model structure, particularly the explicit aggregation of anatomical sites in our framework and the treatment of asymptomatic infections as a single infectious class.

#### 2 PrEP coverage

Table S5: Country-specific estimates of PrEP coverage $\omega$ used for model calibration.

| Country | $P$ | $M$ | $\omega$ |
|---|---|---|---|
| Austria | 2,181 | 3,040,710 | 0.159 |
| Belgium | 8,727 | 3,797,975 | 0.511 |
| Croatia | 686 | 1,225,498 | 0.124 |
| Cyprus | 22 | 319,192 | 0.015 |
| Czechia | 1,874 | 3,515,128 | 0.118 |
| Denmark | 5,193 | 1,913,511 | 0.603 |
| France | 59,326 | 20,878,323 | 0.631 |
| Germany | 40,000 | 26,953,460 | 0.330 |
| Greece | 26 | 3,323,324 | 0.002 |
| Iceland | 300 | 131,573 | 0.507 |
| Ireland | 6,126 | 1,735,856 | 0.784 |
| Italy | 16,222 | 18,817,253 | 0.192 |
| Lithuania | 20 | 943,388 | 0.005 |
| Luxembourg | 681 | 237,948 | 0.636 |
| Malta | 267 | 214,812 | 0.276 |
| Netherlands | 11,934 | 5,812,299 | 0.456 |
| Norway | 3,453 | 1,840,181 | 0.417 |
| Portugal | 6,931 | 3,279,197 | 0.470 |
| Romania | 211 | 6,192,512 | 0.008 |
| Slovenia | 550 | 704,823 | 0.173 |
| Spain | 34,247 | 16,191,803 | 0.470 |
| Sweden | 4,246 | 3,365,681 | 0.280 |

To estimate PrEP coverage, denoted by $\omega$, we used data from the ECDC Evidence Brief Progress towards reaching the Sustainable Development Goals related to HIV in the European Union and European Economic Area: Monitoring implementation of the Dublin Declaration on partnership to fight HIV/AIDS in Europe and Central Asia – 2025 progress report (European Centre for Disease Prevention and Control (2025b); Figure 2), which reports the number of individuals aged 15 years and older who received PrEP at least once during the previous 12 months in 2024 across 22 European countries. We denote this quantity by $P$.

Demographic data were obtained from Eurostat and used to estimate the number of males aged 15–64 years in each country, denoted by $M$. This age range was chosen because it corresponds to the population most relevant to HIV transmission dynamics among men who have sex with men (MSM). Following a common assumption in the literature, we assumed that 3% of males aged 15–64 years are MSM (Marcus et al., 2013). Furthermore, we assumed that a proportion $\alpha = 15\%$ of MSM belong to the high-activity group. The PrEP coverage parameter $\omega$ was therefore estimated as

$$\omega = \frac{P}{0.03 \times M \times \alpha}.$$

Country-specific estimates of $\omega$ are reported in Table S5. To obtain a single representative value for use in the model, we selected the median of the country-specific estimates, i.e., $\omega = 0.31$, rather than the mean, i.e., $\omega = 0.33$. Indeed, the median is less sensitive to extreme values and therefore provides a more robust measure of the central tendency of PrEP coverage across countries.

### 3 Compartmental transmission model

#### 3.1 General case

##### 3.1.1 System of ODEs

$$
\begin{cases}
\dfrac{\mathrm{d}i_{\mathrm{L,S}}}{\mathrm{d}t} = \sigma\Big(1-\alpha-i_{\mathrm{L,S}}-i_{\mathrm{L,A}}\Big)\lambda_{\mathrm{L}} - (\nu+\tau_{\mathrm{S}})i_{\mathrm{L,S}}\,, \\
\dfrac{\mathrm{d}i_{\mathrm{L,A}}}{\mathrm{d}t} = (1-\sigma)\Big(1-\alpha-i_{\mathrm{L,S}}-i_{\mathrm{L,A}}\Big)\lambda_{\mathrm{L}} - (\nu+\tau_{\mathrm{L,A}})i_{\mathrm{L,A}}\,, \\
\dfrac{\mathrm{d}i_{\mathrm{P,S}}}{\mathrm{d}t} = \sigma\Big(\alpha\omega-i_{\mathrm{P,S}}-i_{\mathrm{P,A}}\Big)\lambda_{\mathrm{P}} - (\nu+\tau_{\mathrm{S}})i_{\mathrm{P,S}}\,, \\
\dfrac{\mathrm{d}i_{\mathrm{P,A}}}{\mathrm{d}t} = (1-\sigma)\Big(\alpha\omega-i_{\mathrm{P,S}}-i_{\mathrm{P,A}}\Big)\lambda_{\mathrm{P}} - (\nu+\tau_{\mathrm{P,A}})i_{\mathrm{P,A}}\,, \\
\dfrac{\mathrm{d}i_{\mathrm{U,S}}}{\mathrm{d}t} = \sigma\Big(\alpha(1-\omega)-i_{\mathrm{U,S}}-i_{\mathrm{U,A}}\Big)\lambda_{\mathrm{U}} - (\nu+\tau_{\mathrm{S}})i_{\mathrm{U,S}}\,, \\
\dfrac{\mathrm{d}i_{\mathrm{U,A}}}{\mathrm{d}t} = (1-\sigma)\Big(\alpha(1-\omega)-i_{\mathrm{U,S}}-i_{\mathrm{U,A}}\Big)\lambda_{\mathrm{U}} - (\nu+\tau_{\mathrm{U,A}})i_{\mathrm{U,A}}\,.
\end{cases}
\tag{S1}
$$

##### 3.1.2 Forces of infection

$$
\begin{cases}
\lambda_{\mathrm{L}} &= \pi_{\mathrm{L}}\left(\rho_{\mathrm{LL}}\beta_{\mathrm{LL}}\dfrac{\delta i_{\mathrm{L,S}}+i_{\mathrm{L,A}}}{1-\alpha} + \rho_{\mathrm{LP}}\beta_{\mathrm{LP}}\dfrac{\delta i_{\mathrm{P,S}}+i_{\mathrm{P,A}}}{\alpha\omega} + \rho_{\mathrm{LU}}\beta_{\mathrm{LU}}\dfrac{\delta i_{\mathrm{U,S}}+i_{\mathrm{U,A}}}{\alpha(1-\omega)}\right)\,, \\
\lambda_{\mathrm{P}} &= \pi_{\mathrm{P}}\left(\rho_{\mathrm{PL}}\beta_{\mathrm{PL}}\dfrac{\delta i_{\mathrm{L,S}}+i_{\mathrm{L,A}}}{1-\alpha} + \rho_{\mathrm{PP}}\beta_{\mathrm{PP}}\dfrac{\delta i_{\mathrm{P,S}}+i_{\mathrm{P,A}}}{\alpha\omega} + \rho_{\mathrm{PU}}\beta_{\mathrm{PU}}\dfrac{\delta i_{\mathrm{U,S}}+i_{\mathrm{U,A}}}{\alpha(1-\omega)}\right)\,, \\
\lambda_{\mathrm{U}} &= \pi_{\mathrm{U}}\left(\rho_{\mathrm{UL}}\beta_{\mathrm{UL}}\dfrac{\delta i_{\mathrm{L,S}}+i_{\mathrm{L,A}}}{1-\alpha} + \rho_{\mathrm{UP}}\beta_{\mathrm{UP}}\dfrac{\delta i_{\mathrm{P,S}}+i_{\mathrm{P,A}}}{\alpha\omega} + \rho_{\mathrm{UU}}\beta_{\mathrm{UU}}\dfrac{\delta i_{\mathrm{U,S}}+i_{\mathrm{U,A}}}{\alpha(1-\omega)}\right)\,.
\end{cases}
$$

##### 3.1.3 Sexual mixing matrix

$$
\begin{cases}
\rho_{\mathrm{LL}} &= \dfrac{(1-\alpha)\pi_{\mathrm{L}}+\alpha\omega\epsilon\pi_{\mathrm{P}}+\alpha(1-\omega)\epsilon\pi_{\mathrm{U}}}{(1-\alpha)\pi_{\mathrm{L}}+\alpha\omega\pi_{\mathrm{P}}+\alpha(1-\omega)\pi_{\mathrm{U}}}\,, \\
\rho_{\mathrm{PP}} &= \dfrac{(1-\alpha)\epsilon\pi_{\mathrm{L}}+\alpha\omega\pi_{\mathrm{P}}+\alpha(1-\omega)\epsilon\pi_{\mathrm{U}}}{(1-\alpha)\pi_{\mathrm{L}}+\alpha\omega\pi_{\mathrm{P}}+\alpha(1-\omega)\pi_{\mathrm{U}}}\,, \\
\rho_{\mathrm{UU}} &= \dfrac{(1-\alpha)\epsilon\pi_{\mathrm{L}}+\alpha\omega\epsilon\pi_{\mathrm{P}}+\alpha(1-\omega)\pi_{\mathrm{U}}}{(1-\alpha)\pi_{\mathrm{L}}+\alpha\omega\pi_{\mathrm{P}}+\alpha(1-\omega)\pi_{\mathrm{U}}}\,, \\
\rho_{\mathrm{PL}} &= \rho_{\mathrm{UL}} = \dfrac{(1-\alpha)(1-\epsilon)\pi_{\mathrm{L}}}{(1-\alpha)\pi_{\mathrm{L}}+\alpha\omega\pi_{\mathrm{P}}+\alpha(1-\omega)\pi_{\mathrm{U}}}\,, \\
\rho_{\mathrm{LP}} &= \rho_{\mathrm{UP}} = \dfrac{\alpha\omega(1-\epsilon)\pi_{\mathrm{P}}}{(1-\alpha)\pi_{\mathrm{L}}+\alpha\omega\pi_{\mathrm{P}}+\alpha(1-\omega)\pi_{\mathrm{U}}}\,, \\
\rho_{\mathrm{LU}} &= \rho_{\mathrm{PU}} = \dfrac{\alpha(1-\omega)(1-\epsilon)\pi_{\mathrm{U}}}{(1-\alpha)\pi_{\mathrm{L}}+\alpha\omega\pi_{\mathrm{P}}+\alpha(1-\omega)\pi_{\mathrm{U}}}\,.
\end{cases}
\tag{S2}
$$

#### 3.2 Assortative case – $\epsilon = 1$

##### 3.2.1 System of ODEs

$$\begin{cases} \dfrac{\mathrm{d}i_{\mathrm{L,S}}}{\mathrm{d}t} = \sigma\Big(1-\alpha-i_{\mathrm{L,S}}-i_{\mathrm{L,A}}\Big)\pi_{\mathrm{L}}\beta_{\mathrm{L}}\dfrac{\delta i_{\mathrm{L,S}}+i_{\mathrm{L,A}}}{1-\alpha} - (\nu+\tau_{\mathrm{S}})i_{\mathrm{L,S}}\,, \\ \dfrac{\mathrm{d}i_{\mathrm{L,A}}}{\mathrm{d}t} = (1-\sigma)\Big(1-\alpha-i_{\mathrm{L,S}}-i_{\mathrm{L,A}}\Big)\pi_{\mathrm{L}}\beta_{\mathrm{L}}\dfrac{\delta i_{\mathrm{L,S}}+i_{\mathrm{L,A}}}{1-\alpha} - (\nu+\tau_{\mathrm{L,A}})i_{\mathrm{L,A}}\,, \\ \dfrac{\mathrm{d}i_{\mathrm{P,S}}}{\mathrm{d}t} = \sigma\Big(\alpha\omega-i_{\mathrm{P,S}}-i_{\mathrm{P,A}}\Big)\pi_{\mathrm{P}}\beta_{\mathrm{P}}\dfrac{\delta i_{\mathrm{P,S}}+i_{\mathrm{P,A}}}{\alpha\omega} - (\nu+\tau_{\mathrm{S}})i_{\mathrm{P,S}}\,, \\ \dfrac{\mathrm{d}i_{\mathrm{P,A}}}{\mathrm{d}t} = (1-\sigma)\Big(\alpha\omega-i_{\mathrm{P,S}}-i_{\mathrm{P,A}}\Big)\pi_{\mathrm{P}}\beta_{\mathrm{P}}\dfrac{\delta i_{\mathrm{P,S}}+i_{\mathrm{P,A}}}{\alpha\omega} - (\nu+\tau_{\mathrm{P,A}})i_{\mathrm{P,A}}\,, \\ \dfrac{\mathrm{d}i_{\mathrm{U,S}}}{\mathrm{d}t} = \sigma\Big(\alpha(1-\omega)-i_{\mathrm{U,S}}-i_{\mathrm{U,A}}\Big)\pi_{\mathrm{U}}\beta_{\mathrm{U}}\dfrac{\delta i_{\mathrm{U,S}}+i_{\mathrm{U,A}}}{\alpha(1-\omega)} - (\nu+\tau_{\mathrm{S}})i_{\mathrm{U,S}}\,, \\ \dfrac{\mathrm{d}i_{\mathrm{U,A}}}{\mathrm{d}t} = (1-\sigma)\Big(\alpha(1-\omega)-i_{\mathrm{U,S}}-i_{\mathrm{U,A}}\Big)\pi_{\mathrm{U}}\beta_{\mathrm{U}}\dfrac{\delta i_{\mathrm{U,S}}+i_{\mathrm{U,A}}}{\alpha(1-\omega)} - (\nu+\tau_{\mathrm{U,A}})i_{\mathrm{U,A}}\,. \end{cases} \tag{S3}$$

##### 3.2.2 Endemic prevalence

$$i^*_{k,\mathrm{S}} = n_k\frac{\sigma(\nu+\tau_{k,\mathrm{A}})}{\sigma(\nu+\tau_{k,\mathrm{A}})+(1-\sigma)(\nu+\tau_{\mathrm{S}})}\left(1-\frac{1}{R_{0,k}}\right)\,, \tag{S4}$$

$$i^*_{k,\mathrm{A}} = n_k\frac{(1-\sigma)(\nu+\tau_{\mathrm{S}})}{\sigma(\nu+\tau_{k,\mathrm{A}})+(1-\sigma)(\nu+\tau_{\mathrm{S}})}\left(1-\frac{1}{R_{0,k}}\right)\,. \tag{S5}$$

The total endemic prevalence is

$$i^*_k = i^*_{k,\mathrm{S}} + i^*_{k,\mathrm{A}} = n_k\left(1-\frac{1}{R_{0,k}}\right)\,, \tag{S6}$$

where

$$R_{0,k} = \frac{\pi_k\beta_k(\delta\sigma(\nu+\tau_{k,\mathrm{A}})+(1-\sigma)(\nu+\tau_{\mathrm{S}}))}{(\nu+\tau_{k,\mathrm{A}})(\nu+\tau_{\mathrm{S}})} = \pi_k\beta_k\mathcal{T}_k\,,$$

and

$$n_{\mathrm{L}} = 1-\alpha\,,\ \ n_{\mathrm{U}} = \alpha(1-\omega)\,,\ \text{and}\ n_{\mathrm{P}} = \alpha\omega\,.$$

Note that $\mathcal{T}_k$, which denotes the infectious duration for individuals in group $k$, is given by

$$\mathcal{T}_k = \frac{\gamma\sigma}{\nu+\tau_{\mathrm{S}}} + \frac{1-\sigma}{\nu+\tau_{k,\mathrm{A}}}\,.$$

##### 3.2.3 Endemic incidence

$$\mathcal{I}^*_{k,\mathrm{S}} = (\nu+\tau_{\mathrm{S}})i^*_{k,\mathrm{S}} = \sigma n_k\frac{(\nu+\tau_{k,\mathrm{A}})(\nu+\tau_{\mathrm{S}})}{\sigma(\nu+\tau_{k,\mathrm{A}})+(1-\sigma)(\nu+\tau_{\mathrm{S}})}\left(1-\frac{1}{R_{0,k}}\right) = \frac{\sigma n_k}{\mathcal{D}_k}\left(1-\frac{1}{R_{0,k}}\right)\,, \tag{S7}$$

$$\mathcal{I}^*_{k,\mathrm{A}} = (\nu+\tau_{k,\mathrm{A}})i^*_{k,\mathrm{A}} = (1-\sigma)n_k\frac{(\nu+\tau_{k,\mathrm{A}})(\nu+\tau_{\mathrm{S}})}{\sigma(\nu+\tau_{k,\mathrm{A}})+(1-\sigma)(\nu+\tau_{\mathrm{S}})}\left(1-\frac{1}{R_{0,k}}\right) = \frac{(1-\sigma)n_k}{\mathcal{D}_k}\left(1-\frac{1}{R_{0,k}}\right)\,. \tag{S8}$$

The total endemic incidence is

$$\mathcal{I}^*_k = \mathcal{I}^*_{k,\mathrm{S}} + \mathcal{I}^*_{k,\mathrm{A}} = n_k\frac{(\nu+\tau_{k,\mathrm{A}})\tau_{k,\mathrm{S}}}{\sigma(\nu+\tau_{k,\mathrm{A}})+(1-\sigma)\tau_{k,\mathrm{S}}}\left(1-\frac{1}{R_{0,k}}\right) = \frac{n_k}{\mathcal{D}_k}\left(1-\frac{1}{R_{0,k}}\right)\,. \tag{S9}$$

Note that $\mathcal{D}_k$, which denotes the infection duration for individuals in group $k$, is given by

$$\mathcal{D}_k = \frac{\sigma}{\nu + \tau_{\mathrm{S}}} + \frac{1-\sigma}{\nu + \tau_{k,\mathrm{A}}} \,.$$

##### 3.2.4 Notification rate

$$\mathcal{N}^*_{k,\mathrm{S}} = \tau_{\mathrm{S}} i^*_{k,\mathrm{S}} = \sigma n_k \frac{(\nu + \tau_{k,\mathrm{A}})\tau_{\mathrm{S}}}{\sigma(\nu + \tau_{k,\mathrm{A}}) + (1-\sigma)(\nu + \tau_{\mathrm{S}})} \left(1 - \frac{1}{R_{0,k}}\right) = \sigma n_k \frac{\phi_k}{\mathcal{D}_k} \left(1 - \frac{1}{R_{0,k}}\right) , \tag{S10}$$

$$\mathcal{N}^*_{k,\mathrm{A}} = \tau_{k,\mathrm{A}} i^*_{k,\mathrm{A}} = (1-\sigma) n_k \frac{\tau_{k,\mathrm{A}}(\nu + \tau_{\mathrm{S}})}{\sigma(\nu + \tau_{k,\mathrm{A}}) + (1-\sigma)(\nu + \tau_{\mathrm{S}})} \left(1 - \frac{1}{R_{0,k}}\right) = (1-\sigma) n_k \frac{\phi_k}{\mathcal{D}_k} \left(1 - \frac{1}{R_{0,k}}\right) . \tag{S11}$$

The total notification rate is

$$\mathcal{N}^*_k = \mathcal{N}^*_{k,\mathrm{S}} + \mathcal{N}^*_{k,\mathrm{A}} = n_k \frac{\phi_k}{\mathcal{D}_k} \left(1 - \frac{1}{R_{0,k}}\right) . \tag{S12}$$

Note that $\phi D_k$, which denotes the probability that an infection is detected before natural clearance for individuals in group $k$, is given by

$$\phi_k = \frac{\sigma \tau_{\mathrm{S}}}{\nu + \tau_{\mathrm{S}}} + \frac{(1-\sigma)\tau_{k,\mathrm{A}}}{\nu + \tau_{k,\mathrm{A}}} \,.$$

### 4 Sensitivity of metrics to testing strategies

#### 4.1 Prevalence

For the sake of simplicity, let us assume that $\gamma = 1$, meaning that symptomatic individuals keep having sexual activity, so that both the infection and infectious durations are equal ($\mathcal{T} = \mathcal{D}$). As a reminder, decreasing the asymptomatic testing rate $\tau_{\mathrm{A}}$ lengthens the infection duration. Thus, in order to assess the impact of testing strategies on prevalence, one can compute $\partial i^* / \partial \mathcal{D}$, which satisfies

$$\frac{\partial i^*}{\partial \mathcal{D}} = \frac{1}{\pi \beta \mathcal{D}^2} = \frac{1 - i^*}{\mathcal{D}} \, .$$

The previous equation is always positive, showing that any increase in the infection duration pushes prevalence up.

#### 4.2 Incidence

Let us apply the same reasoning to incidence. In order to assess the impact of testing strategies on incidence, one can compute $\partial \mathcal{I}^* / \partial \mathcal{D}$, which satisfies

$$\frac{\partial \mathcal{I}^*}{\partial \mathcal{D}} = \frac{2 - \pi \beta \mathcal{D}}{\pi \beta \mathcal{D}^3} = \frac{1 - 2i^*}{\mathcal{D}^2} \, .$$

As opposed to prevalence, the previous equation can be either negative or positive. Specifically, it is equal to 0 if and only if $\mathcal{D} = 2/(\pi\beta)$.

### 5 Sensitivity analysis

#### 5.1 High-activity fraction, partner-change rate, and PrEP coverage

In the main text, the high-activity MSM group was defined using a cutoff of $> 20$ partners per year. To assess the sensitivity of our findings to this specific threshold, we tested alternative cutoffs for the distinction between the high- and low-activity groups, specifically $> 10$ and $> 50$ partners per year.

As explained in Section 1.3 and 2, modifying this cutoff directly impacts the estimated fraction of high-activity MSM, $\alpha$, the mean number of partners in both the high- and low-activity groups, $\pi_{\mathrm{H}}$ and $\pi_{\mathrm{L}}$, and the resulting PrEP coverage, $\omega$.

| **Population-specific parameters** | | | | | |
|---|---|---|---|---|---|
| Params. | Meaning | – | >10 | >20 | >50 |
| $\alpha$ | Fraction of high-activity MSM | – | 0.27 | 0.15 | 0.05 |
| $\gamma$ | Symptom-induced activity modification | – | 0.26 | 0.26 | 0.26 |
| $\varepsilon$ | Sexual mixing coefficient | – | 0.59 | 0.64 | 0.67 |
| $\tau_{\mathrm{S}}$ | Symptomatic treatment rate | – | 4.59 $\mathrm{yr}^{-1}$ | 4.31 $\mathrm{yr}^{-1}$ | 4.61 $\mathrm{yr}^{-1}$ |
| $\omega$ | PrEP coverage in high-activity MSM | – | 0.17 | 0.31 | 0.92 |
| **Group-specific parameters** | | | | | |
| Params. | Meaning | Group | >10 | >20 | >50 |
| $\pi$ | Mean number of partners | L | 3.3 $\mathrm{yr}^{-1}$ | 5.2 $\mathrm{yr}^{-1}$ | 8.5 $\mathrm{yr}^{-1}$ |
| | | H | 43.4 $\mathrm{yr}^{-1}$ | 65.3 $\mathrm{yr}^{-1}$ | 128.2 $\mathrm{yr}^{-1}$ |
| $\beta$ | Transmission probability | L | 0.31 | 0.25 | 0.19 |
| | | H | 0.12 | 0.10 | 0.08 |
| $\tau_{\mathrm{A}}$ | Asymptomatic testing rate | L | 0.24 $\mathrm{yr}^{-1}$ | 0.35 $\mathrm{yr}^{-1}$ | 0.60 $\mathrm{yr}^{-1}$ |
| | | H | 0.24 $\mathrm{yr}^{-1}$ | 0.35 $\mathrm{yr}^{-1}$ | 0.60 $\mathrm{yr}^{-1}$ |
| **Disease-specific parameters** | | | | | |
| Params. | Meaning | STI | >10 | >20 | >50 |
| $\sigma$ | Fraction of symptomatic infections | NG | 0.08 | 0.13 | 0.29 |
| $\nu$ | Spontaneous clearance rate | NG | 3.60 $\mathrm{yr}^{-1}$ | 3.94 $\mathrm{yr}^{-1}$ | 4.11 $\mathrm{yr}^{-1}$ |

Table S6: Parameter values for different cutoffs defining the high-activity group.

Using these alternative parameterizations, we ran the ABC calibration procedure again. The parameter values corresponding to each cutoff are summarized in Table S6. We plotted the corresponding heatmaps for prevalence, incidence, and notification rate. As reported by Figure S8, the results remained qualitatively the same across all tested cutoffs, demonstrating that our findings are highly robust to the choice of the threshold used to define the high-activity group.

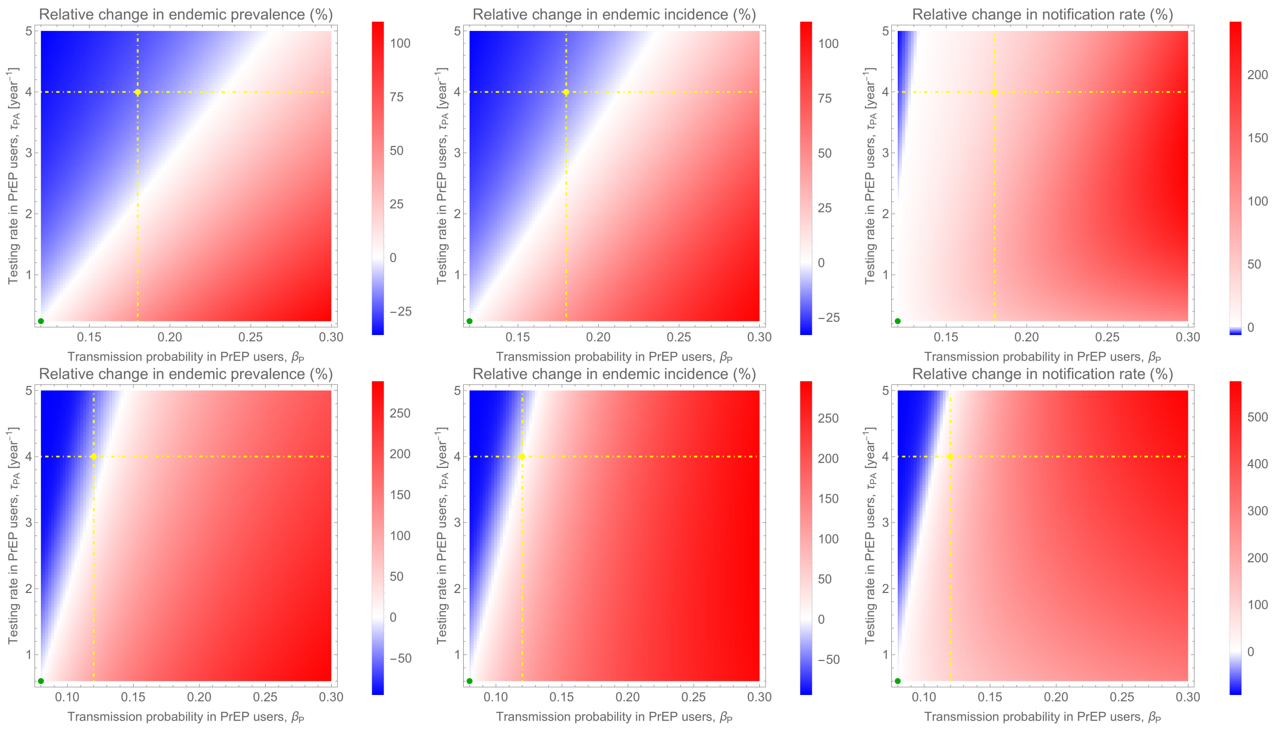

Figure S8: **Sensitivity of epidemiological metrics to alternative sexual activity cutoffs.** The left, middle, and right panels show the relative change in the endemic prevalence, incidence, and notification rate of *Neisseria gonorrhoeae*, respectively, as functions of the asymptomatic testing rate, $\tau_{\mathrm{P,A}}$, and transmission probability, $\beta_{\mathrm{P}}$, among PrEP users. The upper row displays results when the high-activity group is defined by a cutoff of $> 10$ partners per year, while the lower row corresponds to a cutoff of $> 50$ partners per year. Blue regions indicate a decrease in the respective metric, red regions indicate an increase, and white represents no change. The horizontal dot-dashed line corresponds to the standard recommended testing frequency of $\tau_{\mathrm{P,A}} = 4$ year$^{-1}$, and the vertical dot-dashed line highlights a 50% increase in transmission probability. The green point represents the baseline scenario with no behavioral change or differential testing, such that high-activity PrEP users (P) have identical parameter values to high-activity non-users (U). Other parameter values are given in Table S6.

#### 5.2 Threshold transmission probability for PrEP users

Focusing on our cutoff of $> 20$ partners, we further assessed the robustness of our results by analyzing the behavioral conditions required to maintain epidemiological metrics unchanged following the introduction of PrEP.

For each parameter set satisfying the conditions of the ABC calibration, we determined the threshold value of the transmission probability for PrEP users $\beta_{\mathrm{P}}$ that would result in no net change in prevalence, incidence, and notification rate after PrEP introduction. In this analysis, the asymptomatic testing rate for PrEP users was fixed at $\tau_{\mathrm{P,A}} = 4$ year$^{-1}$, as this testing frequency represents the standard recommended care for PrEP users and is well-established.

Figure S9 confirms that our main results are robust. We found that a highly elevated $\beta_{\mathrm{P}}$ value would be necessary for PrEP to cause an actual increase in prevalence and incidence. In contrast, a relatively low $\beta_{\mathrm{P}}$ value is sufficient to induce an increase in the notification rate.

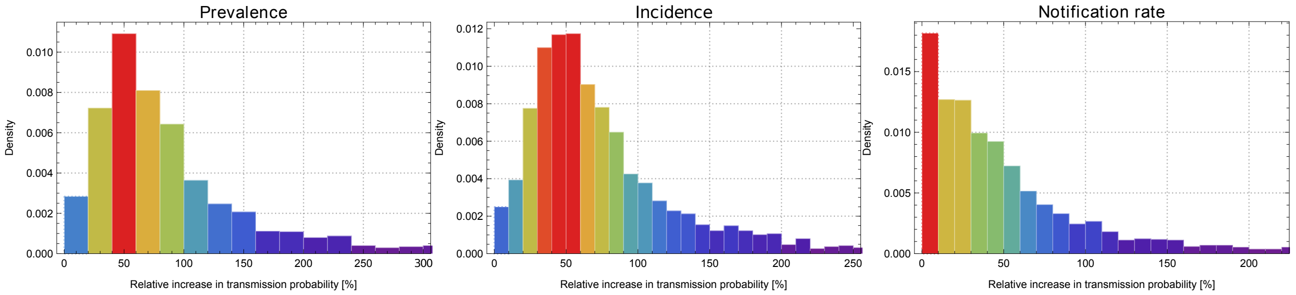

Figure S9: **Distribution of the threshold transmission probability for PrEP users.** The left, middle, and right panels display the distributions of the threshold transmission probability $\beta_P$, expressed as a relative change, required to result in no net change in the prevalence, incidence, and notification rate of *Neisseria gonorrhoeae*, respectively, following the introduction of PrEP. This threshold represents the specific increase in $\beta_P$ at which PrEP introduction has a neutral impact on the corresponding epidemiological metric, assuming PrEP users undergo asymptomatic screening at a frequency of $\tau_{P,A} = 4$ year$^{-1}$. The distributions were generated by computing this threshold for each of the parameter sets retained from the Approximate Bayesian Computation (ABC) calibration procedure.
